# Acceptability, usability and performance of lateral flow immunoassay tests for SARS-CoV-2 antibodies: REACT-2 study of self-testing in non-healthcare key workers

**DOI:** 10.1101/2021.06.21.21259254

**Authors:** Bethan Davies, Marzieh Araghi, Maya Moshe, He Gao, Kimberly Bennet, Jordan Jenkins, Christina Atchison, Ara Darzi, Deborah Ashby, Steven Riley, Wendy Barclay, Paul Elliott, Helen Ward, Graham Cooke

## Abstract

**Background:** Seroprevalence studies in key worker populations are essential to understand the epidemiology of SARS-CoV-2. Various technologies, including laboratory assays and point-of-care self-tests, are available for antibody testing. The interpretation of seroprevalence studies requires comparative data on the performance of antibody tests.

**Methods:** In June 2020, current and former members of the UK Police forces and Fire service performed a self-test lateral flow immunoassay (LFIA) and provided a saliva sample, nasopharyngeal swab, venous blood samples for Abbott ELISA and had a nurse performed LFIA. We present the prevalence of PCR positivity and antibodies to SARS-CoV-2 in this cohort following the first wave of infection in England; the acceptability and usability of self-test LFIAs (defined as use of the LFIA kit and provision of a valid result, respectively); and determine the sensitivity and specificity of LFIAs compared to laboratory ELISAs.

**Results:** In this cohort of non-healthcare key workers, 7.4% (396/5,348; 95% CI, 6.7-8.1) were antibody positive. Seroprevalence was 8.9% (6.9-11.4) in those under 40 years, 11.5% (8.8-15.0) in those of non-white British ethnicity and 7.8% (7.1-8.7) in those currently working. The self-test LFIA had an acceptability of 97.7% and a usability of 90.0%. There was substantial agreement between within-participant LFIA results (kappa 0.80; 0.77-0.83). The LFIAs (self-test and nurse-performed) had a similar performance: compared to ELISA, sensitivity was 82.1% (77.7-86.0) self-test and 76.4% (71.9-80.5) nurse-performed with specificity of 97.8% (97.3-98.2) and 98.5% (98.1-98.8) respectively.

**Conclusion:** A greater proportion of the non-healthcare key worker cohort showed evidence of previous infection with SARS-CoV-2 than the general population at 6.0% (5.8-6.1) following the first wave in England. The high acceptability and usability reported by participants and the similar performance of self-test and nurse-performed LFIAs indicate that the self-test LFIA is fit for purpose for home-testing in occupational and community prevalence studies.

## INTRODUCTION

During the response to the COVID-19 pandemic in the United Kingdom (UK), people who, due to the nature of their work, were unable to work from home were known as “key workers” [1]. The UK key worker population is estimated to be 10.6 million people (33% of the total workforce) with the majority (69%) working in industries other than health and social care, including education and childcare, key public services, transport industry and food sector [1, 2]. It is well established that key workers are at increased risk from infection by SARS-CoV-2 and subsequent COVID-19 [3, 4].

In April 2020 the UK Government initiated a programme of COVID-19 testing which included studies of the prevalence of antibodies to SARS-CoV-2 in the community [5]. In order to facilitate widespread community testing, the REal Time Assessment of Community Transmission-2 (REACT-2) study undertook a programme of development work using self-test finger prick lateral flow immunoassays (LFIAs) for the detection of antibodies to SARS-CoV-2. The initial laboratory validation and usability of the test was carried out among healthcare professionals [6], and further usability and acceptability studies were conducted among a random sample of adults in the population [7].

Here, as part of the REACT-2 programme [8], we aimed both to quantify the prevalence of SARS-CoV-2 (current and past infection) among a subset of public-facing non-healthcare key workers following the first wave of infection in England, and to test at scale the performance of self-test LFIAs versus nurse-administered LFIAs. This was done to determine the acceptability, usability and performance of self-test point-of-care LFIAs in key worker occupations who do not have specialist training in the use of medical devices. Finally, we evaluated the performance of the LFIA against an established ELISA antibody test in sera.

## METHODS

### Cross-sectional study design

Non-healthcare key worker participants were identified through an established occupational research cohort (Airwave Health Monitoring Study), a longitudinal occupational cohort of over 50,000 people who work, or have worked, for the Police forces in the UK [9] and a register of current employees of the West Midlands (UK) Fire Service. All participants with a residential address within a reasonable distance (approximately 50 kilometres) of one of six testing sites (Bournemouth, Derby, Keele, London, Manchester and Warwick) and no known medical condition that might increase bleeding risk were sent a letter or email inviting them to take part together with a participant information sheet. Those who did not respond were sent a reminder at 4 weeks and recruitment was capped at 5,500 participants [8].

### Study procedures

Participants were recruited between 1 June and 10 July 2020. Social distancing (except during finger-prick and venepuncture) and personal protective equipment were implemented at the clinics. Consent was obtained from participants who had been symptom-free for the previous 7-days and they were provided with instructions, a set of study ID stickers (barcode), a self-test LFIA kit, a nasopharyngeal kit and a saliva kit.

Participants were then invited to conduct the LFIA (“self-test”) by following the written/visual instructions provided [10]. No direct instruction was provided from staff at the clinic facility. The LFIA (Fortress, Northern Ireland) used tests antibody against the spike (“S”) protein. It was previously evaluated as having sensitivity 84% (95% CI, 70.5%, 93.5%) and specificity 98.6% (97.1%, 99.4%) [6]. While participants were waiting the 10-15 minutes to read the LFIA test they performed a self-administered saliva collection (2ml, using Oragene kit [DNA Genotek, Canada] and drool technique) using the instructions provided. After reading the LFIA result, participants uploaded an image of the completed self-test LFIA. Participants then underwent a repeat finger-prick LFIA administered by a health care professional (“nurse-performed”) and provided a set of venous blood samples. After leaving the building, participants completed a self-administered nasopharyngeal swab in an outside shelter area or their private vehicle, following the instructions and video, and then placed the sample in the “drop box” at reception. Participants were also asked to complete a brief questionnaire, which included questions on demographic characteristics, occupation, health status (including COVID-19), usability and acceptability of the LFIA device [7].

Samples were transported daily via courier to laboratories for processing. The saliva sample and nasopharyngeal swab were analysed by RT-PCR for SARS-CoV-2 and participants were contacted (email or phone) with the results within three days and, if positive, provided with the latest government instructions on self- and household isolation. Participants had a venous blood sample analysed by ELISA (Abbott, USA) for SARS-CoV-2 IgG antibody. We applied the manufacture recommended diagnostic cut-off (binding ratio of 1.4) to determine a positive result from this quantitative immunoassay [11]. Participants received the results of the blood tests within 4 weeks via letter.

We define the acceptability of the self-test LFIA as the proportion of participants attending the test centre who consented to, and used, the provided kit [7]. We define the usability of the self-test LFIA as the proportion of participants that used the kit who achieved a valid result. A positive LFIA test was defined as IgG positive, and a negative test was defined as IgG negative, regardless of IgM result [7]. For those that did not complete the test, we present the reason given.

### Data analysis

We excluded people without valid questionnaire responses from relevant analyses and retained tests with a positive or negative result. We estimated the prevalence of RT-PCR positivity from nasopharyngeal swab and saliva samples, and antibody positivity was defined as a positive Abbott ELISA test on serum.

After excluding LFIAs with an invalid result, we report the concordance of self-test to nurse-performed LFIA results using the Cohen’s kappa statistic: <0, poor agreement; 0.00–0.20, slight agreement; 0.21–0.40, fair agreement; 0.41–0.60, moderate agreement; 0.61–0.80, substantial agreement; and >0.8, almost perfect agreement [12]. For participants with a valid LFIA (participant and nurse) and Abbott ELISA result, we determined the positive (sensitivity) and negative (specificity) percent agreement of the LFIA tests compared to the serum Abbott ELISA test.

Finally, we undertook an independent visual inspection of the uploaded images from the discordant LFIA test pairs (self-test and nurse-performed) and a similar-sized sample of concordant pairs, to investigate reasons for any differences. Two reviewers (BD and MA) independently reviewed the discordant images and a third (HW) was consulted to adjudicate on and resolve any differences. One reviewer (BD) reviewed the concordant images.

We used logistic regression to quantify associations between demographic characteristics and i) detected antibody status (positive Abbott ELISA test) and ii) not having a valid self-test LFIA result. 95% confidence intervals were calculated using the Wilson’s method. Analyses were conducted in STATA version 13.1 or higher (StataCorp, College Station, TX, USA).

### Ethical approvals

This work was undertaken as part of the REACT-2 study, with ethical approval from South Central–Berkshire B Research Ethics Committee (REC ref: 20/SC/0206; IRAS 283805). Airwave study participants have given consent to be contacted for other research studies (IRAS project ID: 259978).

## RESULTS

Study recruitment was capped at 5,500, therefore of the 24,182 individuals invited to the participate in the study, 5,554 (23.0%) booked into clinic and of these 5,453 (98.2%) attended (Fig. 1). The majority (88%, n=4,729) of participants were 40 years and older, 65% (n=3,483) were men, 92% (n=4,882) were white British (Table 1). Overall, 5,306 (97.3%) participants provided a saliva sample for RT-PCR, 5,436 (99.7%) performed a nasopharyngeal swab for RT-PCR and 5,348 (98.1%) provided blood for an Abbott ELISA test.

**Figure 1.**
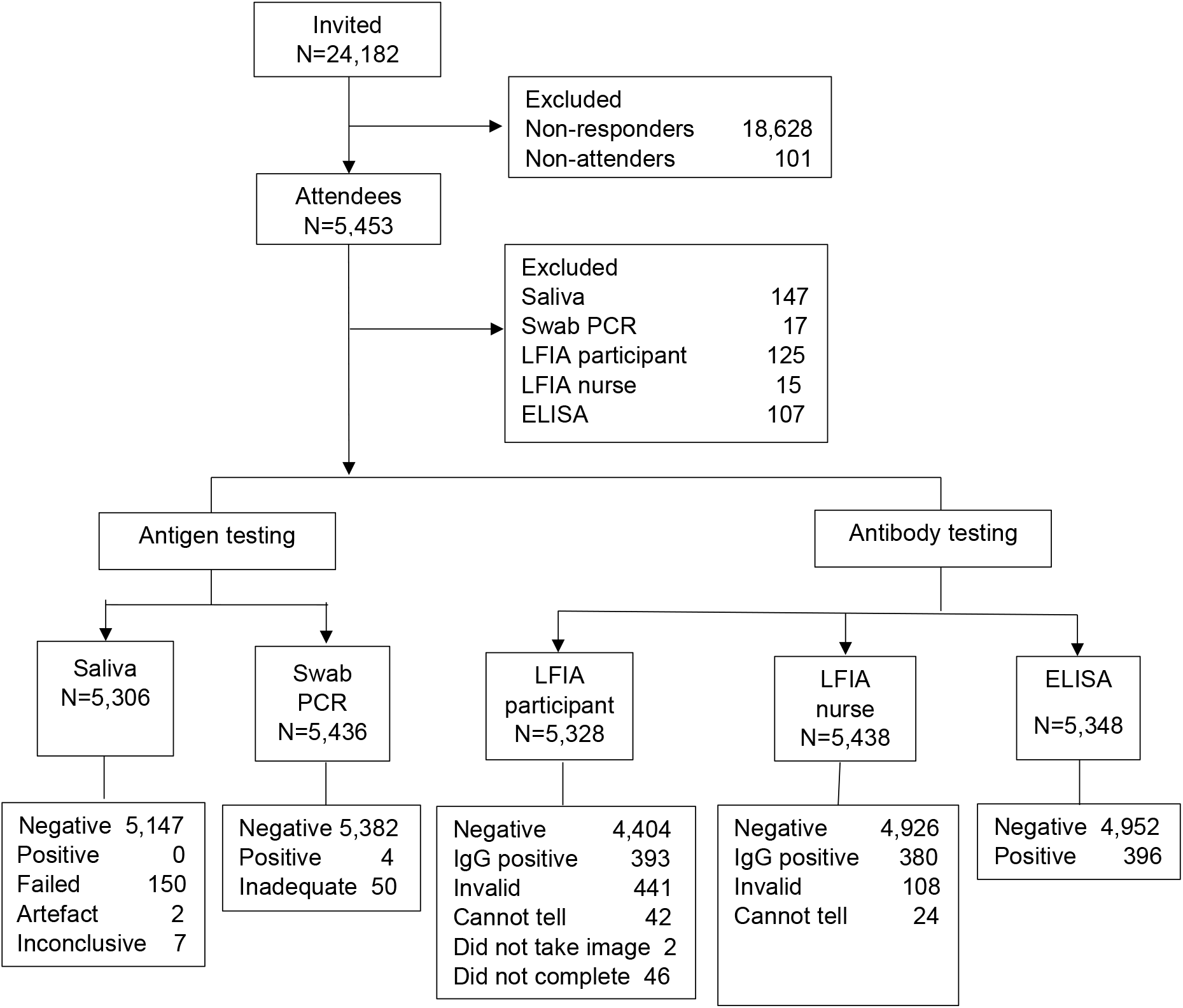
STARD flow diagram.

**Table 1.** Demographic characteristics of participants by antibody status (Abbott ELISA or LFIA) and logistic regression in relation to Abbott ELISA.

|  |  |  | Abbott ELISA |  |  |  |  | LFIA self-test |  |  |
| --- | --- | --- | --- | --- | --- | --- | --- | --- | --- | --- |
| Characteristic |  | Participants<br>N<br>(% of total) | No.<br>positive<br>ELISA/<br>No. valid<br>ELISA<sup>1</sup> | Crude<br>prevalence<br>ELISA<br>% (95% CI) | P<br>value<br><sup>2</sup> | Adjusted OR<sup>3</sup><br>(95% CI) | p<br>value | No.<br>positive/<br>Total<br>valid | Crude<br>prevalence<br>LFIA self-test %<br>(95% CI) | p value |
| Overall |  |  | 396/5348 | 7.4 (6.7- 8.1) |  |  |  | 393/4404 | 8.1 (7.4-9.0) |  |
| Age (years) | <40 | 626 (11.6) | 56/626 | 8.9 (6.9-11.4) | 0.045 | - |  | 62/545 | 11.3 (8.9-14.3) | 0.013 |
|  | 40-49 | 1,686 (31.4) | 125/1684 | 7.4 (6.2-8.7) |  | 0.81 (0.58-1.13) | 0.233 | 122/1497 | 8.1 (6.8-9.6) |  |
|  | 50-59 | 2,192 (40.9) | 170/2189 | 7.7 (6.7-8.9) |  | 0.85 (0.62-1.16) | 0.317 | 152/1935 | 7.8 (6.7-9.1) |  |
|  | 60 and over | 851 (15.8) | 45/849 | 5.3 (3.9-7.0) |  | 0.56 (0.37-0.84) | 0.006 | 47/734 | 6.4 (4.8-8.4) |  |
| Gender | Male | 3483 (65.0) | 266/3478 | 7.6 (6.8-8.5) | 0.354 | - |  | 244/3054 | 7.9 (7.0-9.0) | 0.632 |
|  | Female | 1,872 (34.9) | 130/1870 | 6.9 (5.8-8.1) |  | 0.88 (0.71-1.10) | 0.279 | 139/1657 | 8.3 (7.1-9.8) |  |
| Ethnicity | White | 4882 (92.0) | 336/4789 | 7.0 (6.3-7.7) | 0.001 | - |  | 337/4343 | 7.7 (7.0-8.5) | 0.000 |
|  | Non-white | 424 (7.9) | 48/415 | 11.5 (8.8-15.0) |  | 1.66 (1.20-2.29) | 0.002 | 52/388 | 13.4 (10.3-17.1) |  |
| Currently working | Yes | 4459 (84.2) | 344/4372 | 7.8 (7.1-8.7) | 0.003 | - |  | 346/3992 | 8.6 (7.8-9.5) | 0.012 |
|  | No<sup>4</sup> | 836 (15.7) | 40/821 | 4.8 (3.5-6.5) |  | 0.65 (0.45-0.94) | 0.024 | 43/731 | 5.8 (4.3-7.8) |  |
| Education | Degree | 1629 (30.7) | 109/1600 | 6.8 (5.6-8.1) | 0.299 | - |  | 122/1448 | 8.4 (7.0-9.9) | 0.733 |
|  | No degree | 3678 (69.3) | 275/3605 | 7.6 (6.8-8.5) |  | 1.16 (0.91-1.46) | 0.208 | 267/3284 | 8.1 (7.2-9.1) |  |
| Region | East Midlands | 789 (14.7) | 50/789 | 6.3 (4.8-8.2) | 0.000 | - |  | 46/699 | 6.5 (4.9-8.6) | 0.000 |
|  | London | 1068 (19.9) | 125/1066 | 11.7 (9.9-13.8) |  | 1.90 (1.35-2.68) | 0.000 | 128/956 | 13.4 (11.2-15.6) |  |
|  | North West | 800 (14.9) | 70/799 | 8.7 (6.9-10.9) |  | 1.42 (0.97-2.07) | 0.069 | 58/704 | 8.2 (6.4-10.5) |  |
|  | South West | 764 (14.2) | 23/761 | 3.0 (2.0-4.5) |  | 0.46 (0.27-0.76) | 0.003 | 28/663 | 4.2 (2.9-6.0) |  |
|  | West Midlands | 1,934 (36.1) | 128/1933 | 6.6 (5.5-7.8) |  | 1.06 (0.75-1.48) | 0.732 | 123/1689 | 7.2 (6.1-8.6) |  |

<sup>1</sup>Participants with a valid questionnaire and valid Abbott ELISA result; <sup>2</sup> p-value from Chi squared test; <sup>3</sup>Adjusted for age and sex; <sup>4</sup> includes people in government supported training, unemployed and available for work, wholly retired from work, full-time education at school, college, or University, looking after home/ family, permanently sick / disabled, and “doing something else”.

Four (0.07%; 95% CI, 0.03, 0.19) participants were RT-PCR positive from nasopharyngeal swabs; none of 5,306 saliva tests were positive. Due to the low numbers testing positive, no further analysis of prevalence of SARS-CoV-2 infection is presented.

### Antibody prevalence

IgG antibodies to SARS-CoV-2 were found in 396 of 5,348 participants using the Abbott ELISA, a prevalence of 7.4% (6.7, 8.1). Prevalence varied by age being highest in people under 40 years of age (8.9%; 6.9,11.4) and lowest in people over 60 years (5.3%; 3.9, 7.0); by ethnicity, being higher in non-white (11.5%; 8.8, 15.0) compared to white (7.0%; 6.3, 7.7) participants; employment status, being higher in people currently working (7.8%; 7.1, 8.7) compared to people not currently working (4.8%; 3.5, 6.5) and region: higher in London (11.7%; 9.9, 13.8) compared to other regions except North West and lowest in the South West (3.0%; 2.0, 4.5) (Table 1). These associations persisted after controlling for age and sex, with the odds of positivity in London being almost twice those in East Midlands (Odds Ratio [OR] of 1.90 [1.35, 2.68]), and OR in non-white people of 1.66 (1.20, 2.29) compared to white people (Table 1). Three hundred and ninety-three (8.1%; 7.4, 9.0) participants were antibody positive on the self-test LFIA.

### Comparative performance of antibody tests

For participants with a valid result from self-test LFIA, nurse-performed LFIA and Abbott ELISA, 95.2% (4,363/4,582) were concordant (positive or negative) across all three tests (Supplementary Table S1). Compared to Abbott ELISA, self-test LFIAs had a sensitivity of 82.1% (77.7, 86.0) and specificity of 97.8% (97.3, 98.2) and nurse-performed LFIAs a sensitivity of 76.4% (71.9, 80.5) and a specificity of 98.5% (98.1, 98.8) (Table 2a).

**Table 2.** Comparison of results from (a) LFIAs and Abbott ELISA, and (b) self-test LFIA and nurse-performed LFIA.

| **(a)** |  | **ELISA** |  |  |  |
| --- | --- | --- | --- | --- | --- |
|  |  | **Positive** | **Negative** | **Total** | **Performance (95% CI)** |
| **Self-test LFIA** | **Positive** | 285 | 98 | 383 | Sensitivity: 82.1% (77.7, 86.0) |
|  | **Negative** | 62 | 4260 | 4322 | Specificity: 97.8% (97.3, 98.2) |
|  | **Total** | 347 | 4358 | 4705 |  |
| **Nurse-performed LFIA** | **Positive** | 298 | 72 | 370 | Sensitivity: 76.4% (71.9, 80.5) |
|  | **Negative** | 92 | 4744 | 4836 | Specificity: 98.5% (98.1, 98.8) |
|  | **Total** | 390 | 4816 | 5206 |  |

| **(b)** |  | **Nurse-performed LFIA** |  |  |  |
| --- | --- | --- | --- | --- | --- |
|  |  | **Positive** | **Negative** | **Total** | **% agreement (95% CI)** |
| **Self-test LFIA** | **Positive** | 294 | 93 | 387 | Positive: 88.8% (84.9, 92.0) |
|  | **Negative** | 37 | 4246 | 4283 | Negative: 97.9% (97.4, 98.3) |
|  | **Total** | 331 | 4339 | 4670 | Kappa: 0.80 (0.77, 0.83) |

There was substantial agreement between the two LFIA results (self-test and nurse-performed) from the same participants, with 4,540/4,670 (97.2%) having the same result, kappa 0.80 (0.77, 0.83) (Table 2b). Overall, there were discordant results between the self-test and nurse-performed tests in 130 cases (2.8%). Uploaded images were available for review from 101 of these discordant pairs, 56 (55%) of which were confirmed as discordant (Supplementary Table S2). The review showed that participants were more likely to report negative tests as positive, and nurses were more likely to miss faint positives, which may explain the lower sensitivity of nurse-tests compared to Abbott ELISA (Supplementary Table S3). Review of the uploaded images for the sample of 145 concordant self-test and nurse-performed tests found that 129/129 available image pairs were concordant (Supplementary Table S4).

### Self-test LFIA acceptability and usability

The self-test LFIA test was attempted by 5,328 participants, an acceptability of 97.7%. Of the participants who attempted the test, 4,797 obtained a valid result, giving a usability of 90.0%. The most commonly cited reasons for not successfully completing the LFIA were “I could not read the result” (n=78), “It was too fiddly for me to manage” (n=51) and “I did not manage to get the buffer onto the test” (n=37). An invalid self-test LFIA result was 20% less likely for women compared to men (OR 0.78; 0.63, 0.97) and 40% more likely for people who were not currently working compared to people currently working (OR 1.38; 1.03, 1.85) (Supplementary Table S5). The majority of participants (88.5%, n=4,638) were “very confident” that they had interpreted their results correctly. Of the fifty participants who provided a preference for where to perform the test, 82.0% (62.9, 90.2) stated home.

## DISCUSSION

One in 13 people (7.4%; 6.7, 8.1) in this non-healthcare key worker cohort had evidence of previous infection with SARS-CoV-2 following the first wave of the epidemic in England, rising to one in nine in London (11.7%; 9.9, 13.8). This prevalence was higher than in a representative sample of the population at the time at 6.0% (5.8, 6.1), but lower than in key workers in healthcare at 11.7% (10.5, 13.1) [4]. The participants reported a high acceptability and usability of self-test LFIAs, with 90% of participants obtaining a valid antibody test result. There was no significant difference in the performance of self-test and nurse-performed LFIAs. The prevalence of infection in the cohort was low at 0.07% (n=4) on RT-PCR from nasopharyngeal swab, and none of the saliva samples tested positive. This low prevalence reflects the declining epidemic at the time of the study, during June to July 2020 [13, 14].

The Fortress LFIA used in this study followed a rigorous evaluation of five commercially available LFIAs [6]. We used the Abbott ELISA as the primary reference standard. An evaluation by Public Health England (PHE), found that it met the manufacturer-reported performance for specificity, although not for sensitivity [11].

Our study provides further evidence for the association of occupation, involving key workers in a public-facing role, with risk of infection by SARS-CoV-2. Previous research suggests that in mid-June to mid-July, the crude prevalence in key workers (excluding health and care-home workers) in England was 6.1% (5.7, 6.4) [15], over a percentage point lower than the 7.4% (6.7, 8.1) reported here. However, it is possible that the prevalence following the first wave of the pandemic among participants in our study could be under-estimated given uncertainty about antibody waning and the duration of a detectable antibody response [16]. We found similar demographic patterns in prevalence to those seen in the national REACT-2 study, where the highest prevalence was also found in the London region, in people of non-white ethnicity and younger adults [4].

The sensitivity of self-test LFIAs estimated against Abbott ELISA in this cohort of non-healthcare key workers was 82.1% (77.8, 85.8), similar to previously reported sensitivity of 84% (70.5, 93.5) in a cohort of healthcare workers with PCR-confirmed COVID-19 [6]. The specificity against Abbott ELISA in this study was 97.8% (97.3, 98.2), similar to the 98.6% (97.1, 99.4) using pre-pandemic sera in the laboratory [6].

Overall, there was substantial agreement between self-test to nurse-performed LFIAs, with only 2.8% having discrepant results. There was no evidence that nurse-performed tests were better than self-tests, although there was a suggestion that participants were more likely to record a test as positive and healthcare practitioners were more likely to report as negative. This was supported by the visual reinspection of the small number of discordant pairs of LFIAs. The study suggests that there is no gain in accuracy to be had by committing extra resource to obtain healthcare practitioner performed LFIAs in large-scale community surveys and that self-test is a viable and acceptable approach.

### Strengths and limitations

We aimed to replicate the experience of completing a home self-test LFIA as a means of obtaining prevalence of previous SARS-CoV-2 infection in non-healthcare occupational settings by providing participants with no additional assistance, beyond the test instruction materials. The idea was that, if successful, this could then be extended to obtain prevalence estimates in the wider population at low cost, without the need for supervision by a healthcare practitioner. We found little difference between the results of LFIAs administered as a self-test or by a nurse, supporting the unsupervised use of the LFIA. However, our results might not be fully generalisable. The majority of participants were recruited from the Airwave occupational cohort [9]. The cohort is not demographically (age, sex, ethnicity, geography) representative of the adult population of England, participants had to be able to travel to a test site (mostly in a private vehicle) and, being an occupational cohort, participants were healthier than the general population (‘healthy cohort’ effect [9]). Participants (emergency service staff) may also have been more familiar with medical procedures, which may have had an impact on the generalisability of the usability and acceptability findings to the general population. Nonetheless, parallel studies carried out in the community at around the same time provided similar results on usability and acceptability (although estimates of prevalence were lower, reflecting greater exposure of public-facing workers to risk of infection) [7].

We compared results of the self-test and nurse-performed LFIA with a quantitative ELISA test (Abbott) using an established cut-point to denote positivity. To the extent that the “gold standard” ELISA test itself does not have perfect sensitivity and specificity [11], will have introduced some error into those comparisons.

## CONCLUSION

In June 2020, following the first wave of the COVID-19 pandemic in England, approximately one in 13 of this non-healthcare key worker cohort had evidence of past infection with SARS-CoV-2. The epidemiological patterns in prevalence of previous infection were similar to the national picture, with highest prevalence observed in people who were under 40 years, had a non-white ethnicity, were currently in employment and living in London. Participants reported a high acceptability and usability for the LFIAs and there was little difference in performance of self-test compared to a nurse-performed LFIA. Overall, our study suggests that the self-test LFIA is fit for purpose for home-testing for use in occupational and – by extension – community prevalence studies of anti-SARS-CoV-2 antibodies.

## Data Availability

Aggregate data from study participants is presented in tables and supplementary information.

## DATA AVAILABILITY

Aggregate data from study participants is presented in tables and supplementary information.

## FUNDING

This work was funded by the Department of Health and Social Care in England. Authors acknowledge funding from National Institute of Health Research (NIHR) Professorship [to G.C.], NIHR Senior Investigator Award [to A.D., H.W.], Medical Research Council Centre for Environment and Health [MR/L01341X/1, MR/S019669/1 to P.E.],NIHR Imperial College NHS Trust Biomedical Research Centre [to P.E, H.W.], NIHR Applied Research Collaborative [to H.W.], NIHR Health Protection Research Units in Chemical and Radiation Threats and Hazards [to P.E., B.D.] and in Environmental Exposures and Health [to P.E.], British Heart Foundation Centre for Research Excellence at Imperial College London [RE/18/4/34215 to P.E.], Wellcome Trust [200861/Z/16/Z, 200187/Z/15/Z to H.W.], Health Data Research UK (HDR UK) [to P.E.], and UK Dementia Research Institute at Imperial [MC_PC_17114 to P.E.]. We thank The Huo Family Foundation for their support of our work on COVID-19.

## ACKNOWLEDGEMENTS

We thank key collaborators on this research: Department of Epidemiology and Biostatistics at Imperial College: Eric Johnson and Rob Elliott; Institute of Global Health Innovation at Imperial College: Gianluca Fontana, Sutha Satkunarajah and Lenny Naar; Airwave study: Andy Heard, Paul Downey, Antoinette Amuzu.

## DECLARATION OF INTERESTS

AD is Chair of the Health Security initiative at Flagship Pioneering UK Ltd.. All other authors declare no competing interests.

## Notes

### Competing Interest Statement

AD reports as Chair of the Health Security initiative at Flagship Pioneering UK Ltd.. All other authors declare no competing interests.

### Clinical Protocols

https://doi.org/10.12688/wellcomeopenres.16228.2

### Author Declarations

This work was undertaken as part of the REACT-2 study, with ethical approval from South Central Berkshire B Research Ethics Committee (REC ref: 20/SC/0206; IRAS 283805). Airwave study participants have given consent to be contacted for other research studies (IRAS project ID: 259978).

### Summary of Updates

Conflict of interest statement corrected.

