## Supplementary data for "Acceptability, usability and performance of lateral flow immunoassay tests for SARS-CoV-2 antibodies: REACT-2 study of self-testing in non-healthcare key workers" (from another version of this preprint: version 1, 2021-06-25)

### Supplementary Materials

#### Comparative performance of antibody tests

Comparison was made of self-test LFIA, nurse-performed LFIA and Abbott ELISA where valid test results were available for all three tests. The large majority (95.2%, 4,363/4,582) was concordant (positive or negative) across all three tests (Supplementary Table S1).

Visual inspection of the photographs (jpeg images) of the LFIAs, taken and uploaded at the time of the test, was performed to identify reasons for discordant results. Of the 130 pairs of LFIAs with discordant results, there were 127 self-test and 129 nurse-performed images available for review. Two reviewers independently recorded the result of the test and a third reviewer was consulted for images where consensus was not reached.

After excluding a single unreadable nurse image (blurred) and the images with an invalid result, there were 101 pairs available for review (Supplementary Table S2). Of these, 56 pairs were confirmed as discordant by the reviewers, i.e. the discordant results were confirmed rather than based on misreading of a result. This equates to 56 of 4670 (1.2%) test pairs, indicating good reproducibility (98.8%) of the test on the same day.

Self-test results were reclassified more often than nurse-performed results (46% (n=58/127) compared to 20% (n=25/128) (Supplementary Table S3). Participants were more likely to incorrectly score negative results as positive, while nurses were more likely to incorrectly report positive results as negative.

A further visual inspection of a sample of concordant self-test and nurse-performed LFIA pairs was performed. For each of the six testing sites we selected: eleven pairs of negative tests (defined as IgG and IgM negative) by taking every other person from first entry; the first eleven pairs of positive tests (IgG or IgG+IgM) and all pairs of IgM positive tests (i.e. negative but not in the first category). After excluding unreadable images (participant n=5, nurse n=3) and missing images (participant n=8) there were 129 pairs available for review (Supplementary Table S4). Of these, all 129 pairs were confirmed as concordant by the reviewer.

**Supplementary Table S1.** Summary of participant test results for cohort with three valid test results.

| **Self-test LFIA** | **Nurse-performed LFIA** | **ELISA** | **Number of participants** |
|---|---|---|---|
| Negative | Negative | Negative | 4,123 |
| Positive | Positive | Positive | 240 |
| Negative | Negative | Positive | 46 |
| Negative | Positive | Negative | 20 |
| Negative | Positive | Positive | 16 |
| Positive | Negative | Negative | 49 |
| Positive | Negative | Positive | 41 |
| Positive | Positive | Negative | 47 |
| **TOTAL** | | | **4582** |

**Supplementary Table S2.** Summary of reviewer result for the pairs of LFIAs discordant on initial nurse/participant assessment and with two images of LFIAs with a valid result.

| | | Nurse-performed | | |
|---|---|---|---|---|
| Self-test | | Positive | Negative | Total |
| | Positive | 22 | 38 | 60 |
| | Negative | 18 | 23 | 41 |
| | Total | 40 | 61 | 101 |

**Supplementary Table S3**. Comparison of original rater's assessment with consensus assessment from reviewers; (a) participant, (b) nurse.

a) Participant

| | | Reviewer result n (column %) | | | |
|---|---|---|---|---|---|
| Participant rated self-test | | Positive | Negative | Invalid | Total |
| | Positive | 51 (81.0) | 27 (60.0) | 12 (63.2) | 90 |
| | Negative | 12 (19.0) | 18 (40.0) | 7 (36.8) | 37 |
| | Total | 63 | 45 | 19 | 127 |

b) Nurse

| | | Reviewer result n (column %) | | | |
|---|---|---|---|---|---|
| Nurse rated nurse- performed | | Positive | Negative | Invalid | Total |
| | Positive | 31 (63.3) | 2 (2.7) | 4 (80.0) | 37 |
| | Negative | 18 (36.7) | 72 (97.3) | 1 (20.0) | 91 |
| | Total | 49 | 74 | 5 | 128 |

**Supplementary Table S4.** Summary of reviewer result for the pairs of LFIAs concordant on initial nurse/participant assessment and with two images of LFIAs with a valid result.

| | | Nurse-performed | | |
|---|---|---|---|---|
| Self-test | | Positive | Negative | Total |
| | Positive | 68 | 0 | 68 |
| | Negative | 0 | 61 | 61 |
| | Total | 68 | 61 | 129 |

**Supplementary Table S5.** Logistic regression of the factors associated with having an invalid self-test LFIA test result.

| Characteristic | | No. of participants[1] (%) | Usability (No. with invalid result) | Proportion invalid result, % (95% CI) | p value | aOR[2] (95%CI) | p value |
|---|---|---|---|---|---|---|---|
| **Overall** | | 5328 | 441 | | | | |
| **Age (years)** | <40 | 604 (11.7) | 59 | 9.7 (7.6-12.4) | 0.559 | - | |
| | 40-49 | 1632 (31.7) | 135 | 8.2 (7.0-9.7) | | 0.83 (0.60-1.15) | 0.280 |
| | 50-59 | 2103 (40.8) | 168 | 7.9 (6.9-9.2) | | 0.79 (0.57-1.08) | 0.140 |
| | 60 and over | 804 (15.6) | 70 | 8.7 (6.9-10.8) | | 0.86 (0.59-1.23) | 0.418 |
| **Gender** | Male | 3356 (65.2) | 302 | 8.9 (8.0-10.0) | 0.034 | - | |
| | Female | 1787 (34.7) | 130 | 7.2 (6.1-8.5) | | 0.78 (0.63-0.97) | 0.031 |
| **Ethnicity** | White | 4749 (92.0) | 406 | 8.5 (7.7-9.3) | 0.079 | - | |
| | Non-white | 413 (8.0) | 25 | 6.0 (4.1-8.8) | | 0.65 (0.43-1.01) | 0.056 |
| **Occupation status** | Currently working | 4342 (84.2) | 350 | 8.0 (7.2-8.9) | 0.110 | - | |
| | Not currently working[3] | 810 (15.7) | 79 | 9.7 (7.8-11.9) | | 1.38 (1.03-1.85) | 0.027 |
| **Education** | Degree | 1595 (30.8) | 147 | 9.2 (7.8-10.7) | 0.131 | | |
| | No degree | 3568 (69.1) | 284 | 7.9 (7.1-8.8) | | 0.83 (0.67-1.02) | 0.090 |
| **Region** | East Midlands | 767(14.9) | 68 | 8.8 (7.0-11.0) | 0.132 | | |
| | London | 1027(20.0) | 71 | 6.9 (5.5-8.6) | | 0.74 (0.52-1.06) | 0.104 |
| | North West | 763 (14.8) | 59 | 7.7 (6.0-9.8) | | 0.87 (0.60-1.25) | 0.458 |
| | South West | 719 (14.0) | 56 | 7.7 (6.0-9.9) | | 0.87 (0.60-1.26) | 0.470 |
| | West Midlands | 1867 (36.3) | 178 | 9.5 (8.2-10.9) | | 1.09 (0.81-1.46) | 0.542 |

[1] *Excludes "cannot tell" (n=42), "did not take photo" (n=2), "did not complete the test" (n=46);* [2] *Adjusted for age and sex;* [3] *includes people in government supported training, unemployed and available for work, wholly retired from work, full-time education at school, college, or University, looking after home/ family, permanently sick / disabled, and "doing something else".*
